# Single-Cell Transcriptomics of Multi-Site Cell Therapy in Osteoarthritis: Tissue-Specific Traits and Treatment Correlations

**DOI:** 10.1101/2025.04.14.25325743

**Authors:** Paramita Chatterjee, Hazel Y. Stevens, Linda E. Kippner, Carolyn Yeago, Hicham Drissi, Kenneth Mautner, Scott Boden, Greg Gibson, Krishnendu Roy

## Abstract

Knee-Osteoarthritis (Knee-OA) is a prevalent joint disorder lacking FDA-approved cell-therapies to halt its progression. This study uses single-cell RNA sequencing to analyze bone marrow aspirate concentrate (BMAC) and stromal vascular fraction (SVF) samples in a clinical trial of autologous cell therapies. Trial site-specific variability was significant in BMAC, necessitating tailored normalization, whereas SVF was less affected, likely due to uniform subcutaneous fat sampling. Variance partitioning and tensor decomposition identified site effects in BMAC but revealed shared pathways across cell types in both tissues. Differential gene expression (DEG) analysis between responders and non-responders yielded no significant findings, though likelihood ratio testing (LRT) revealed enrichment for DEG patterns linked to disease severity, potentially masked by patient heterogeneity. Key BMAC pathways included oxidative phosphorylation, unfolded protein response, and TNFα signaling. Cell-cell communication analysis suggested enhanced HLA signaling in non-responder MSCs, consistent with inflammation, while responders showed more coordinated immune interactions. BMAC-MSCs promoted chondrocyte proliferation, whereas SVF-MSCs emphasized immune regulation. Variability in therapy outcomes reflects patient heterogeneity beyond genomic factors, complicating the immediate use of genomic profiling to guide treatment. Nonetheless, as molecular pathways are better understood, integrating genomic insights into personalized strategies may become feasible.

## Introduction

Osteoarthritis (OA) is a degenerative joint disease that significantly diminishes the quality of life for millions of people worldwide. It is characterized primarily by debilitating joint pain and progressive cartilage loss^1^. Traditional treatment modalities, such as physical therapy, pharmacological interventions, and intra-articular corticosteroid injections, provide symptomatic relief but often fail to arrest disease progression or restore damaged tissues effectively. In recent years, the emergence of regenerative medicine has improved the prospective therapeutic landscape for OA by introducing innovative cellular therapies designed to exploit the body’s inherent regenerative capabilities^2^. Among these therapies, autologous bone marrow aspirate concentrate (BMAC)^3^ and adipose-derived stromal vascular fraction (SVF)^4^ have gained prominence. BMAC is rich in stromal cells that aim to harness the regenerative and immunomodulatory properties of bone marrow-derived cells to tackle the multifaceted pathology of OA, which includes cartilage degradation, subchondral bone remodeling, and inflammation. Similarly, SVF^5^, derived from adipose tissue, contains a heterogeneous mix of cells with potent anti-inflammatory and regenerative properties, offering a promising avenue for managing knee OA by mitigating inflammation and promoting repair within the joint.

Despite the promise and growing clinical application of these therapies, a thorough evaluation of their efficacy, particularly in comparison to other cellular therapies and standard of care, remains a critical unmet need^6^. To address this, a comprehensive phase 2/3 multicenter, single-blind, randomized controlled clinical trial, MILES (NCT03818737), was undertaken. This trial aimed to assess the safety and efficacy of BMAC, SVF, and allogeneic human umbilical cord tissue-derived mesenchymal stromal cells against conventional treatments for knee OA in a robust study design involving 480 patients^7^. The trial’s methodology involved randomized allocation of patients into treatment arms receiving cellular therapies or corticosteroid treatments, with follow-up extending to three years. Central to this study was the innovative integration of tissue-specific single-cell RNA sequencing analysis, which was employed to probe the molecular and cellular dynamics within bone marrow and adipose.

Transcriptomic profiling was performed to determine whether there is a correlation between the cellular dynamics observed in treatments like BMAC and clinical outcomes such as pain relief and improved knee function. Association between molecular profiles and patient-reported outcomes enhances our understanding of how cellular therapies modulate the OA environment at a molecular level, potentially leading to more tailored and effective treatment strategies. Our previous study^8^ reported on the use of single-cell transcriptomics to analyze bone marrow aspirate concentrate (BMAC) from OA patients, revealing significant alterations in key cell types, including mesenchymal stromal cells (MSCs), natural killer (NK) cells, monocytes, B cells, and T cells between OA patients and controls. The findings suggested that bone marrow plays a significant role as a reservoir for cellular mediators of inflammation and immune modulation in OA.

The current study expands on this work by incorporating additional BMAC profiles along with data from stromal vascular fractions (SVF) from OA patients and exploring their relationship to therapeutic response. This study thus provides a more detailed characterization of the cellular environments and interactions within these tissues. Overall, the inter-individual variability of profiles obscures any major signatures of remission or disease progression, though we do identify gene expression features that may be enriched in some recipients with poor outcomes. Efforts to incorporate personalized genomic profiling into therapeutic decision making will need to account for the patient variability.

## Materials and Methods

### Study Design and Participants

The MILES^9^ study was a multicenter single-blinded clinical trial performed in accordance with guidelines and oversight from the FDA, and under the management of a contracted research organization. Study approval was obtained from the Western Institutional Review Board and by Duke and Emory University’s institutional review boards. In accordance with the FDA requirements, an investigational new drug application was filed (#18414), referencing investigative device exemption #17894. The study was also registered with ClinicalTrials.gov^7^ (NCT03818737). A total of 570 patients were screened to identify 480 eligible patients who were randomized at five clinical sites across five states within the United States. The five clinical sites included clinics from Emory University (GA), Sanford Health (SD and ND), the Andrews Institute (FL), and Duke University (NC). Participants were randomized to a four-cohort parallel-design study. Participants were further subdivided into three arms, each of which was matched with one third (n=40) of the corticosteroid injection (CSI) controls (**Figure 1A**), arm 1—autologous bone marrow concentrate (BMAC) (n = 120); arm 2—autologous SVF (n = 120); arm 3—allogeneic umbilical cord tissue (UCT) (n = 120). The control cohort of CSI was subsequently aggregated to allow a 1:1:1:1 comparison for analysis. Subjects were enrolled if they were between 40 and 70 years of age and carried a diagnosis of knee OA as determined by radiographs within 3 months of their clinical visit, also having a Kellgren-Lawrence (KL) OA severity score between 2-4. A full list of eligibility criteria is provided on ClinicalTrials.gov (NCT03818737). In the BMAC arm, a total of 12 subjects did not complete the study: 1 related to release criteria, 10 related to subject withdrawal, and 1 lost to follow-up. In the SVF cohort, 12 subjects did not complete the study: 1 due to investigator withdrawal, 4 related to release criteria, 6 related to subject withdrawal, and 1 due to screen failure. In the UCT cohort, 13 subjects did not complete the study: 2 due to investigator withdrawal, 10 related to subject withdrawal, and 1 lost to follow-up. In the CSI cohort, 11 subjects did not complete the study: 1 due to investigator decision, 6 related to subject withdrawal, and 4 lost to follow-up. Of the remaining patients, samples from 134 patients from each of the BMAC and SVF arms, and samples from all 8 UCT-MSC lots met minimum criteria for additional processing and were analyzed for single cell RNAseq (**Figure 1B**). Patients returned to the clinic for MRI to assess cartilage and joint health^10^ at baseline, 6 months, and 1 year.

**Figure 1:**
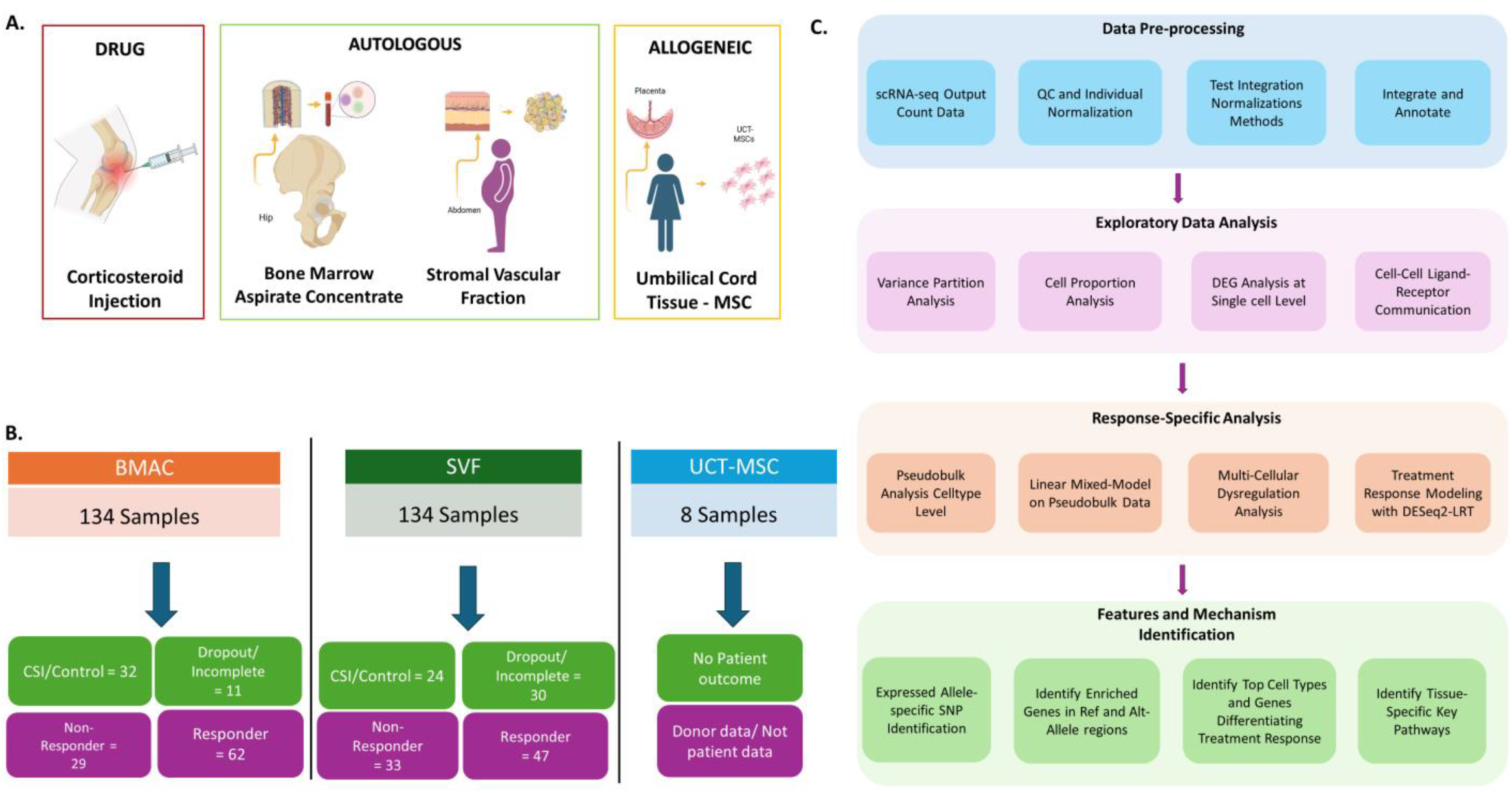
Experimental and Analytical Design and for scRNAseq data. **A.** MILES Clinical Trial Data Collection Design: The trial consisted of 480 total participants, randomized into one of 3 arms. In each arm, patients received either cell therapy injection or corticosteroid injection (control) to the knee. BMAC and SVF cell product consisted of autologous therapeutic cells collected from the patient’s bone marrow or adipose tissue, respectively. The UCT-MSC allogeneic cell therapy was manufactured from umbilical cord tissue, with 8 lots included. **B**. Out of the total enrolled patients in the study, samples from 134 patients from each of the BMAC and SVF arms, and 8 samples from UCT-MSC lots were analyzed using 10x Genomics 3’ scRNAseq kit V3.1. All the autologous data from BMAC and SVF are collected pre-treatment. For BMAC, among 134, there are 32 patients treated with corticosteroid injection (CSI), 11 patients dropped out, 29 patients were grouped as non-responders as of VAS responsiveness and 62 patients were responders as of the VAS responsiveness after 12 months follow-up. For SVF, among 134, there are 24 patients treated with corticosteroid injection (CSI), 30 patients dropped out, 33 patients were grouped as non-responders as of VAS responsiveness and 47 patients were responders as of the VAS responsiveness after 12 months follow-up. For UCT-MSCs being allogeneic, there were data generated from 8 donor samples who are not OA patients. C. Data analysis pipeline design

### Randomization and Masking

Subjects were stratified by clinical center, and after informed consent was obtained and eligibility criteria verified, they were randomized to treatment groups^11^. Treatment assignments were also stratified by clinical center and generated using a pseudo-random number generator with randomly permuted blocks. These assignments were stored in the Medidata Rave cloud-based data management system developed by the contracted research organization. All subjects underwent the harvesting procedure per their assigned study arm, and then, according to the randomization scheme, they were blinded to the actual treatment received (e.g., SVF versus CSI, BMAC versus CSI, or cells versus CSI). As a single-blinded study, the site principal investigators were not required to be blinded; however, physicians and subjects were blinded to the injection. The blinding was implemented by limiting visualization of the syringe contents with opaque coverings, in addition to performing sham BMAC and SVF harvests in patients within certain cohorts.

### Single-Cell RNA Sequencing Analysis

To promote consistency, standardized protocols were followed across the five study sites for cellular harvesting, product preparation, injection procedures, and corticosteroid injections (CSI). While BMAC and SVF samples were fresh autologous products, MSCs derived from umbilical cord tissue had been cryopreserved and prepared to cGMP standards at Duke University. In our earlier publication^8^, 77 BMAC samples from the MILES cohort were used to compare healthy and disease conditions; an additional 57 samples in this study expanded the total to 134, following the same protocols. Single-cell suspensions from cryopreserved BMAC and SVF samples were prepared and assessed using the NC-200 Nucleocounter cell counter. Transcriptome analysis was conducted on both the BMAC and SVF cells, as well as the allogeneic UCT-MSC samples. Library preparation and sequencing were performed using the 10x Genomics Chromium platform with the 3′ V3.1 kit on an Illumina Novaseq 6000. The analysis pipeline is depicted in **Figure 1C**. Raw data were processed with Cell Ranger (10X Genomics) to generate count matrices, which were then analyzed with the Seurat (v4.0) R package^12^. Cells were filtered by quality, requiring 500-10,000 UMIs per cell and <20% mitochondrial gene content, with genes detected in fewer than 3 cells excluded. For BMAC samples, the median cell count per sample was 3680 (s.d. 2218) and the median reads per cell were 45282 (s.d. 86678). For SVF samples, the median cell count per sample was 4869 (s.d. 2058) and the median reads per cell were 105555 (s.d. 3746).

After normalization, principal component analysis (PCA) was used to reduce dimensionality. Among several normalization approaches tested, including robust principal component analysis (rPCA)^13^, canonical correlation analysis (CCA) and Harmony^14^, rPCA was selected for its ability to balance batch effects with biological signal retention. Clustering was performed using the Louvain algorithm, and after quality control comparison we settled on parameters that identified 37 cell clusters.

### Cell Type Annotation and Abundance Assessment

Identified clusters were annotated with cell types based on the expression of known marker genes from literature and databases. The DISCO^15^ database (https://www.immunesinglecell.org/ct_pub) was utilized for automated annotation, supplemented by manual review. For ambiguous clusters, additional marker gene analyses and literature review were performed to assign the most probable cell type. The results were visualized using UMAP^16^, highlighting the distribution and relationship of the annotated cell types within the samples.

A matrix of cell counts per identified cell type for each sample was prepared by aggregating the UMI counts for all genes within a cell, thereby assigning each cell to its annotated cell type based on the clustering and annotation steps previously described. Subsequently, the relative abundance of each cell type across different samples and conditions was quantified using the Propeller tool^17^.

### Pseudobulk Differential Gene Expression Analysis

To investigate the relationship between differential gene expression and clinical response in the trial arms, pseudobulk^18^ profiles were generated with customized scripts by aggregating single-cell RNA-seq counts across annotated cell types within each sample. Initial quality control included principal component analysis (PCA) and hierarchical clustering on log-transformed or variance-stabilized data to assess sample similarity and detect outliers. To mitigate site-specific effects across clinical centers, batch correction was applied to the pseudobulk data using CombatSeq^19^. Identification of differentially expressed genes (DEGs) was then performed using DESeq2^20^. For BMAC, we utilized likelihood ratio tests (LRT)^21^ to compare full and reduced models (excluding age, sex, and BMI) to identify enriched sets of DEGs. In parallel, variance component partitioning was performed with dreamlet^22,23^. For the SVF dataset, in addition to the pseudobulk analyses, single-cell RNA-seq data was also analyzed with scITD which uses Tucker tensor decomposition to capture gene sets co-regulated across multiple cell types. All results were adjusted for multiple comparisons using the Benjamini-Hochberg method with a 5% false discovery rate (FDR)^24^ threshold.

### Cell-to-Cell Communication Analysis

CellChat^25^ and NicheNet^26^ were used to identify likely signaling networks and pathways that may mediate intercellular interactions. CellChat was used to analyze and visualize cell-cell communication networks based on inferred ligand-receptor interactions previously annotated in a comprehensive well-validated database. NicheNet further generates sender-receiver models of cellular communication, prioritizing ligand-target links that are most likely to explain observed gene expression changes in receiver cells. Both tools were implemented with standard workflows recommended by the developers.

### Pathway Enrichment Analysis

Pathway enrichment analysis was conducted using a combination of Gene Set Enrichment Analysis (GSEA)^27^ and its optimized counterpart, fast Gene Set Enrichment Analysis (fgsea)^28^, to rank genes based on their differential expression and assess the enrichment of predefined gene sets. The ESCAPE R^29^ package was employed for the integration of pathway databases, enabling comprehensive visualization and interpretation of enriched pathways. Statistical significance was determined using nominal p-values and adjusted for multiple comparisons also with a 5% FDR threshold.

## Results

### Cluster Analysis Reveals Distinct Cell Type Compositions in BMAC and SVF Datasets

Cluster analysis of the BMAC and SVF datasets revealed 18 and 19 distinct cell-type clusters, respectively (**Figure 2A and 2B**). In the BMAC dataset, major cell types identified included immune cells, monocytes, erythrocytes, megakaryocytes (MEGA), mesenchymal stromal cells (MSCs), and hematopoietic stem cells (HSPCs), with MSCs comprising a low percentage, approximately 0.2% of the total cell population after filtering and quality control. In the SVF dataset, predominant cell types included adipose-derived stromal cells, epithelial cells, fibroblasts, macrophages, and MSC which constituted approximately 5% of the total cell population. The distinction between adipose-derived stromal cells (ASCs) and MSCs was based on marker expression: ASCs exhibited high CD34 and HLA-DR expression but low CD90, while MSCs were characterized by high CD90 and low HLA-DR expression. Following extensive comparison of different procedures for integration of datasets obtained from the different collection sites, we settled on rPCA for all subsequent analyses. As with all single cell RNAseq analyses, the final cluster assignment represents a balance between consistency across datasets, noise reduction, and matching cell types to biological expectation. Erythrocytes were removed from the final clustering as their proportion is highly variable and obscures trends in proportions of the other cell types.

**Figure 2:**
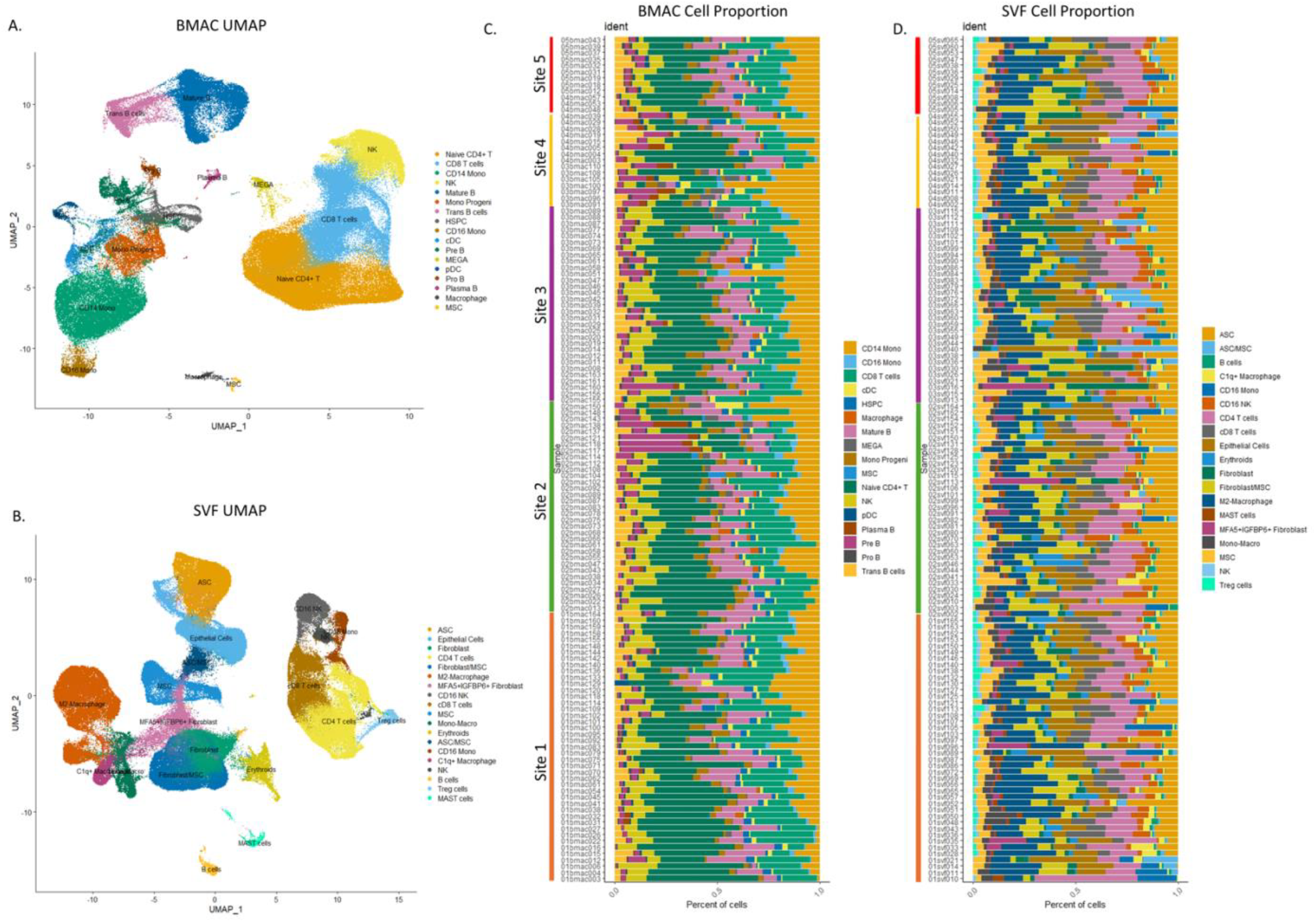
UMAPs and Cellular compositions for scRNAseq celltype clusters for BMAC and SVF datasets. **A**. Celltypes identified for BMAC datasets showing cellular composition. 18 large celltype clusters are identified. The erythrocyte cluster was excluded to reduce the noise for BMAC. Large immune celltypes and monocytes are identified. **B**. BMAC cell composition barplot aligned with clinical collection site. **B**. Celltype identified for SVF datasets showing the cellular composition of SVF as a product. 19 large celltype clusters are identified among these immune cell, epithelial cell, macrophages, adipose derived stromal cells and fibroblasts are identified. **D**. SVF cell composition barplot aligned with clinical collection site. Dittoseq package is used for the barplots, and Propeller tool is used for the statistical analysis on the cell proportions to investigate any significant correlation between cell types and the metadata like Sex, Site, Age, Individuals, and Treatment responses.

### Cell Abundance Analysis

Inferred cell type proportions per donor group by collection site are shown in **Figure 2C, D** for BMAC and SVF respectively. Using the Propellor tool to evaluate variability of cell type abudance, in the BMAC, macrophages, progenitor B and plasma cells, CD14 monocytes, and megakaryocytes were all found to vary among sites regardless of normalization method (**Table S1**). This is most likely due to subtle differences in bone marrow sampling by different physicians. Since pro-B and mature B cells, dendritic cells, MSC as well as naïve CD4 and CD8 T cells were additionally significantly variable among sites when using Harmony, we regarded this method as less robust but show the proportions in **Figure S1**. In contrast, variability across normalization methods was not observed for the SVF datasets shown in **Figure 2D**. After controlling for collection site, age, sex, BMI and Kellgren-Lawrence disease severity index, no cell type proportions varied significantly between responders and non-responders (**Tables S1-S4**).

### Variance Partition Analysis for BMAC and SVF

Turning our attention to gene expression within cell types, dreamlet was used to perform variance partition analysis of the influence of collection site (**Figure 3A**) and therapeutic response status (**Figure 3B**). The violin plots show the range of variance explained by these factors for each expressed gene with the main effect shown in brown and residual unexplained variance in gray. These results imply a substantial influence of collection site on BMAC gene expression variability, with a median between 10% and 25% in most BMAC cell types, for some genes over 50% of the variability due to site, most notably in MSCs and plasma B cells. Only CD16 positive monocytes showed an elevated collection site influence in the SVF dataset.

**Figure 3:**
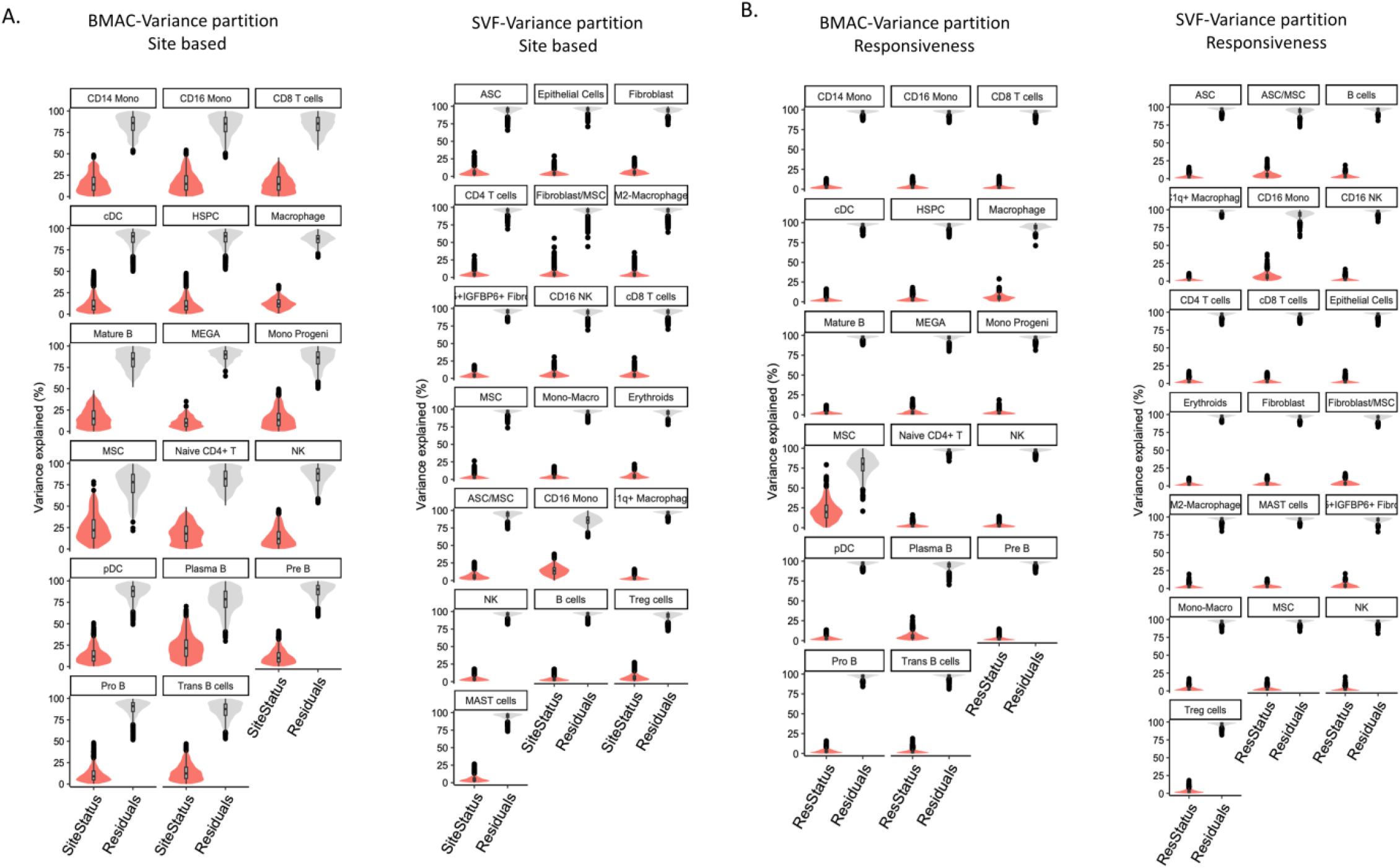
Assessment of site variance impact on gene expression in two separate datasets. **A**. This figure depicts the impact of the site variable on gene expression for various cell types within two independent datasets, visualized side by side for comparison (Left: BMAC, Right: SVF). Violin plots are used to represent the distribution of variance percentages explained by the site variable within each cell type. This plot illustrates BMAC with cell types from CD14 Monocytes to Transitional B cells. The black dots indicate the median variance explained by the site, and the vertical lines delineate the interquartile range for each cell type. The distribution and spread of the violin plots convey the degree to which the site variable influences gene expression within each cell type. A broader plot suggests a greater diversity in the variance explained by the site, whereas a narrower plot indicates less variability. By comparing the range and median values of the variance explained across the two panels, we can infer which dataset exhibits a more substantial effect of the site on gene expression. This analysis employs the VariancePartition from Dreamlet package, which utilizes linear mixed models to quantify the fractional contribution of the site variable to the overall expression variation.

In strong contrast, treatment response contributed minimally to the variance in both BMAC and SVF datasets, explaining less than 10-15% of the variability of all genes across most cell types. Interestingly, the only exception was MSCs in the BMAC with approximately 20% of the variance in gene expression associated with treatment response, though this result is tempered by the relatively small counts of MSC in that tissue compartment. There was also a slightly elevated response of ASC/MSC and CD16 monocyte gene expression in the SVF.

### Identification of Multicellular Gene Expression Programs Across Diverse Cell Types Using scITD in Osteoarthritis SVF: Insights into Donor-Specific Variabilities and Immune-Stromal Interactions

We next performed tensor decomposition to explore how gene expression programs shared across multiple cell types associate with biological or technical aspects of the study. For this, we used scITD software, which provided the most informative results for gene expression programs associated with immune and stromal cell interactions in the SVF dataset. The software first identifies components of variation shared across cell types and shows how these covary with collection site or treatment response (**Figure 4A**) and then allows visualization of the expression of key transcripts associated with the components in the contributing cell types. Factors 2 and 4 weakly associated with site, but none of the first five factors associated with response. Nevertheless, Factor 1 exhibited a strong association with specific immune cell types (**Figure 4B**), notably antibody-secreting cells, B cells, CD4 T cells, NK cells, macrophages, and monocytes. Upregulated genes associated with Factor 1 included *FBN1, IGFBP5*, and *MFAP5*, all potentially engaged in immune-stromal interactions, while *TGFBR3* and *PRG4* were down-regulated in these cell types. Gene Set Enrichment Analysis (GSEA) of the Factor 1 loadings further supported stromal cell enrichment for pathways related to innate immune response, B cell-mediated immunity, and T cell activation. In parallel, pathways linked to cytokine signaling, antigen presentation, and immune effector functions were highlighted in the immune cells in the SVF. Loadings of other gene sets and cell types for Factors 2 through 5 are shown in **Figure S2**.

**Figure 4:**
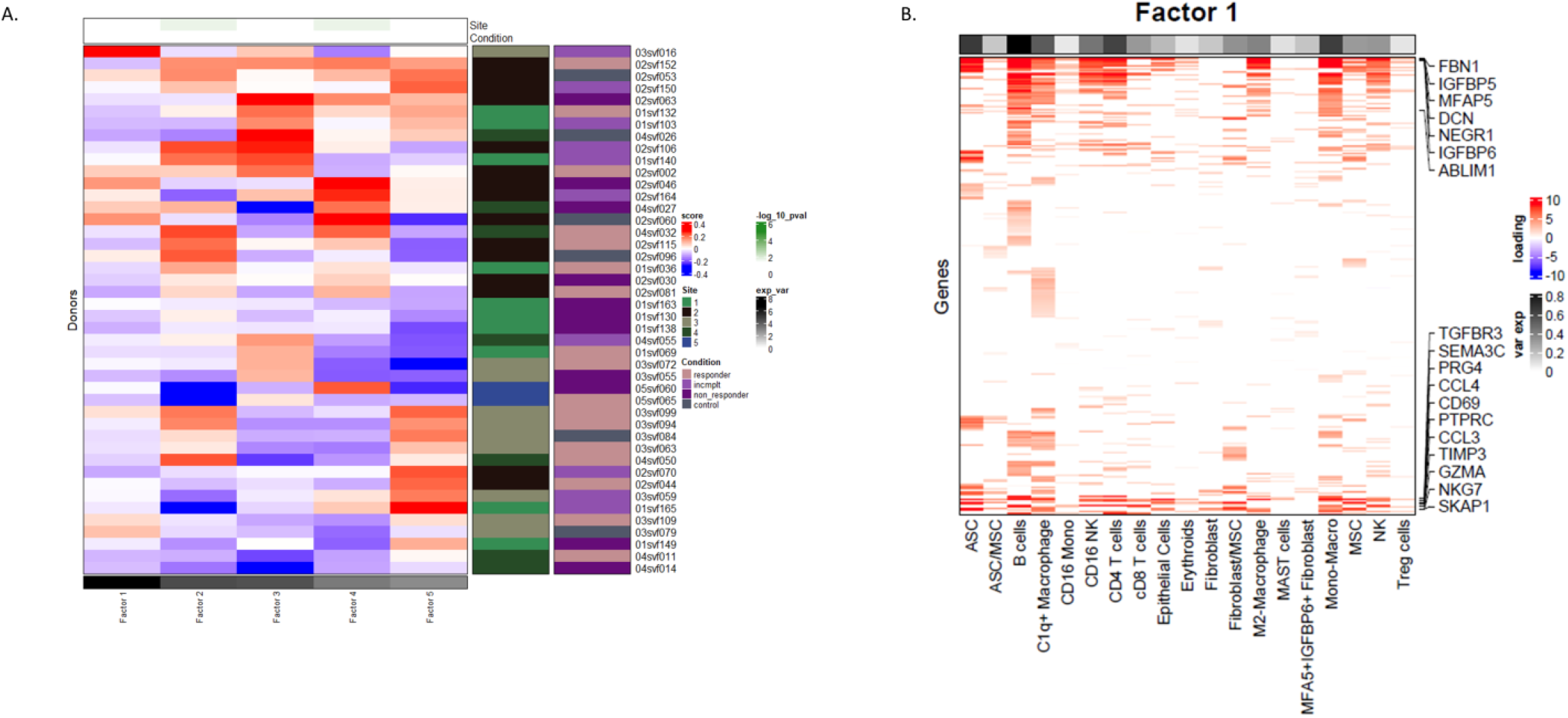
Tensor Factorization of Gene Expression Profiling in SVF. **A**. The heatmap depicts z-score normalized expression data across multiple donors, stratified by site and condition, with sidebars indicating sample origin (Site) and clinical outcome (Condition). Red shades indicate higher factor levels, and blue shades represent lower factor levels relative to the mean. **B**. The heatmap illustrates the association of various genes with ‘Factor 1’ across different cell types. Red intensity signifies a stronger association. Listed genes demonstrate a pronounced association with Factor 1, suggesting a potential biological significance in these specific cell types

### Pseudobulk Analysis of BMAC shows enrichment of DEGs correlated with Clinical Response

Reflecting the above documented strong site effect, scITD Factor 1 for BMAC was strongly associated with collection site. Since the remaining BMAC factors did not clearly associate with a biological covariate, an alternative analytical approach was taken to better address whether response to therapy correlates with differential gene expression. Single cell profiles were converted to pseudobulk profiles for each cell type in each donor. Since there was no significant differential gene expression between responders and non-responders below the 5% false discovery rate threshold in any of the individual cell types after standard hypothesis testing, we utilized the likelihood ratio test (LRT) option in DESeq2 to contrast a null model of no differences among response groups with alternate models. We thus searched for genes whose profile in pseudobulk cells representing each cell type in all individuals was more likely to fit a specific pattern of differential expression with respect to treatment response. **Figure 5A** shows that in all 18 BMAC cell types by far the most commonly supported model showed a gradient from lowest expression in samples from individuals who did not complete the study to highest in responders, who generally resemble control. The number of genes following this pattern ranged from 2,923 in mature B cells (and similarly in the major T cell sub-types) to fewer than 100 in MSC and macrophages (reflecting the small number of such cells).

**Figure 5:**
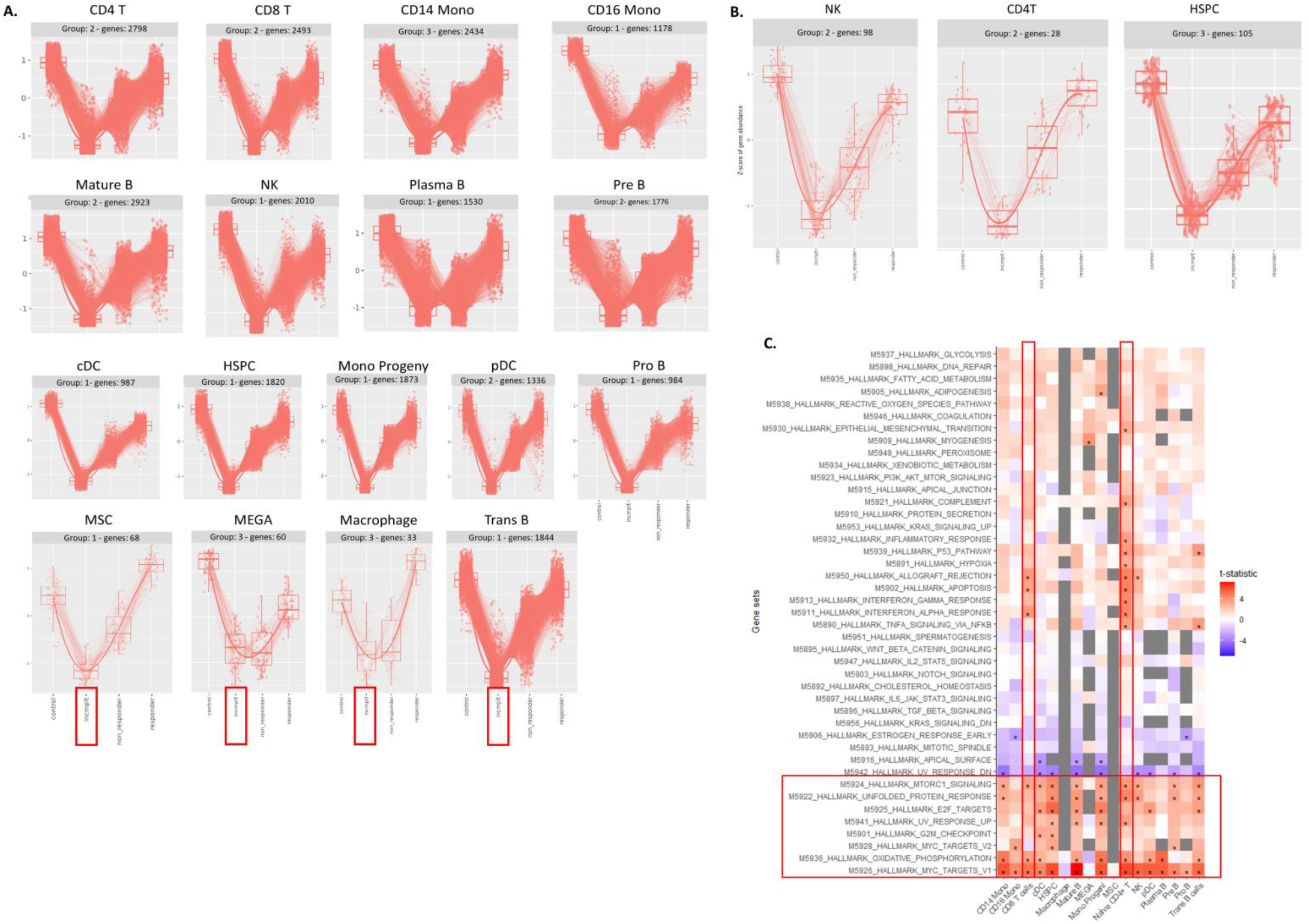
Differential gene expression analysis across treatment groups using DESeq2 likelihood ratio test (LRT). **A.** The plots visualize the z-score of gene abundance for multiple cell type containing groups of DEGs with a consistent pattern. Each subplot represents a distinct group, labeled by group number and the count of genes analyzed. The categories on the x-axis denote the different treatment conditions: control, incomplete responder, non-responder, and responder. Points represent individual genes, and the lines connect the median z-scores across the treatment conditions, showing the relative expression levels. Box plots overlay the data points to summarize the distribution of expression within each condition. The color red signifies genes belonging to ‘one’ category, as indicated by the color legend. **B**. The pattern is consistent after removing the site variability from the data. **C**. Key pathways identified in heatmap showcases various inflammatory and degenerative differential pathways, with significance denoted by asterisks in the non_responders. GSEA database is used for the pathway analysis.

Further adjustment was performed using Combat to remove the batch effect of collection site on the pseudobulk expression profiles. This considerably reduced the numbers of significant genes as much as two orders of magnitude, yet around 100 genes with this pattern were retained, with examples show in **Figure 5B**. Gene set enrichment analysis (GSEA) of these DEGs identified key biological pathways impacted by the treatment. Upregulated pathways included KRAS signaling and hypoxia, while downregulated pathways encompassed oxidative phosphorylation, unfolded protein response, and TNFα signaling via NFκB (**Figure 5C**).

Restricting the analysis to the responder and non-responder categories shows that the lack of significant differential gene expression between groups may be due to the high variability within groups. This is clearly illustrated by the dot plots for the ten most differentially expressed genes in four cell types in **Figure 6 (need to decide if we want to move to supplement or keep in main figures)**. The size of each dot represents the proportion of cells for the donor who express the transcript, and the shading represents the average gene expression. We chose ten individuals of each class at random from across the collection sites, and grouped them by response group, but visually it is clear that there is little correspondence between response status and expression. The four selected cell types—CD16 monocytes, Pro-B cells, Plasma B cells, and MSCs—display variation in gene expression without clear separation between responders and non-responders. In CD16 monocytes, for example, while most of the ten genes are co-expressed at a higher level in non-responders, one responder (labelled 04bmac039) is among the high-expressors, while others have elevated expression of one or a few of the genes. A similar pattern by the other cell types, with the MSCs showing highly variable expression of multiple ribosomal protein-encoding genes. These results indicate that gene expression variation does not distinctly separate responders from non-responders.

**Figure 6:**
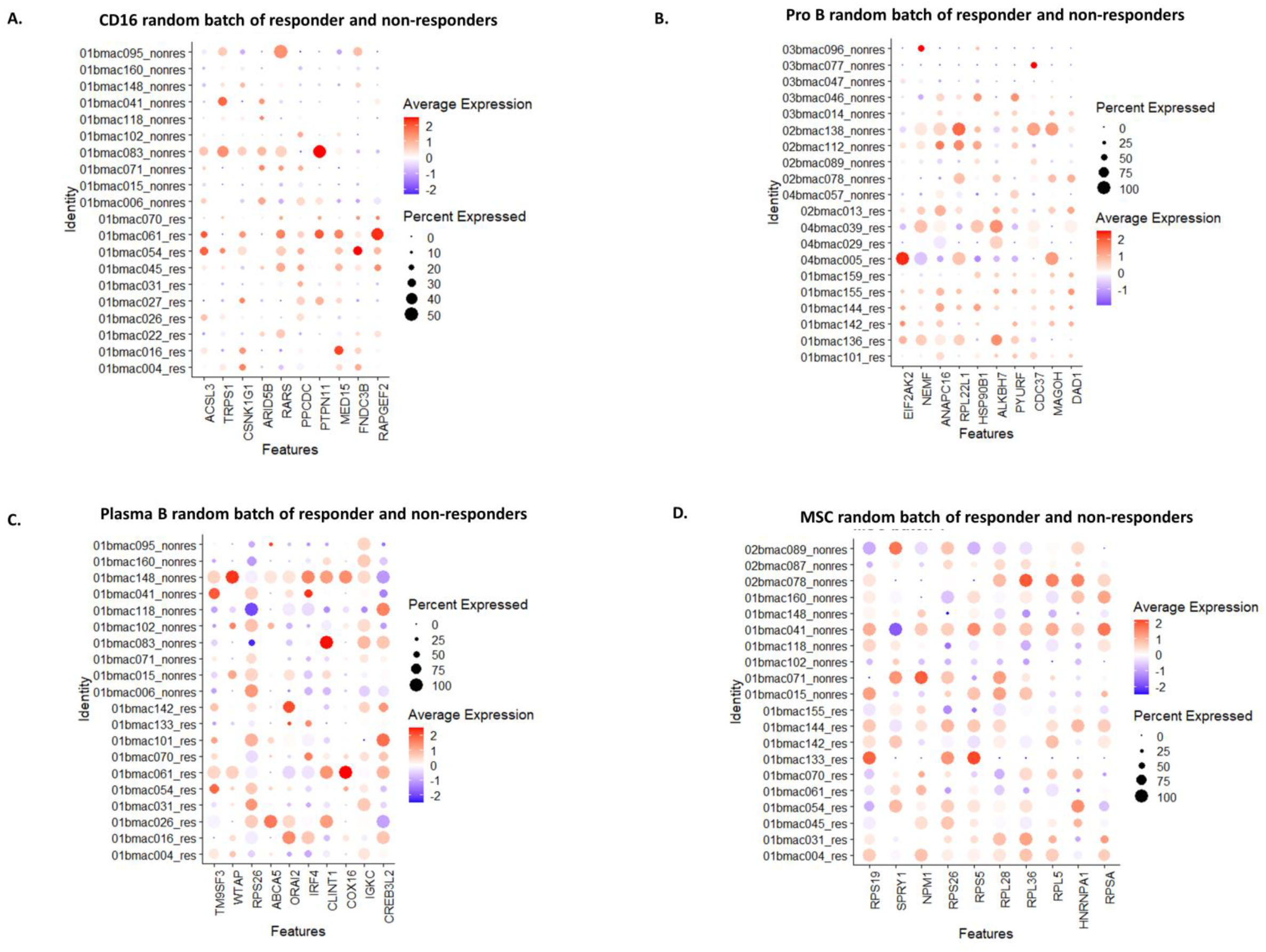
Expression Variability of Selected Genes in BMAC Cell Types Across Random Batches. This figure illustrates the expression variability of the top 10 most differentially expressed genes (DEGs) in selected BMAC cell types across randomly assigned batches of 20 samples. Each dot represents a gene (y-axis) within a specific cell type (x-axis). Dot size reflects the percentage of cells expressing the gene, and color intensity denotes the average expression level, with red indicating higher expression and blue indicating lower expression. **(A-D)** The selected BMAC cell types display expression diversity for the most differentially expressed 10 genes, highlighting variability in gene expression patterns across sample batches.

### Cell-to-Cell Communication Analysis in BMAC and SVF

Cell-to-cell communication analysis was conducted to ask whether there is potential for interactions between mesenchymal stromal cells (MSCs) and other cell types to differ between responders and non-responders. Using the CellChat tool, which infers likely receptor-ligand interaction networks from scRNA-seq data, a rich network of interactions was observed, with MSCs potentially capable of establishing communicative links with every cell type analyzed, and significant autocrine interactions noted in every cell type except macrophages (**Figure S3A**). Focusing on the top ten differentially expressed ligand-receptor interactions (**Figure S3B**), it was found that MSCs were pivotal predicted communicators in responder-derived BMAC cells. The relative information flow in **Figure S3C** suggests that MSCs in responder samples exhibited interactions enriched for ligands such as THY1, CD40, and IGF1, which are associated with immune modulation and tissue repair. By contrast, non-responder samples seem to be enriched for ligands such as IGFBP and CSF that are more typically linked to inflammatory and cellular stress responses.

A similar analysis was performed on the SVF single cell RNAseq dataset, contrasting predicted cell communication differences between non-responders as the reference group and responders as the target condition. This analysis revealed ASC, MSC and M2 macrophages as the most prominent signal sender cell types in SVF (**Figure 7A**). The identities of the ASC and MSC populations, expression of gene markers shown in **Figure 7B** was used to separate ASCs and MSCs based on distinct expression profiles. MSCs exhibit higher expression of *THY1* (CD90) and *ENG*, while ASCs are characterized by reduced *CD34, HLA-DRA*, and *HLA-DRB1* expression. Focusing on specific ligand-receptor interactions and target gene predictions, several ligands enriched in the responder group were identified, both in terms of the number of interactions and their relative strengths. Dot plots in **Figure 7C** illustrate how the expression of various ligands differ between the three cell types. Notably, FGF7 is significantly upregulated in MSCs but is downregulated in ASCs, and conversely, IL15 is downregulated in MSCs and upregulated in ASCs. Other significant ligands enriched in MSCs include *COL5A2, ANGPT1, POSTN, LAMA4*, and *S100A4*, while ligands such as *F8, NRG2*, and *VWF* are upregulated in ASCs. Additionally, the heatmap highlights the differential expression of genes predicted by NicheNet to be targets of the ligands, including *CCL3, CCL4, IL2RB, STAT4, TRAF1, SESN1*, and *NFKB1*.

**Figure 7:**
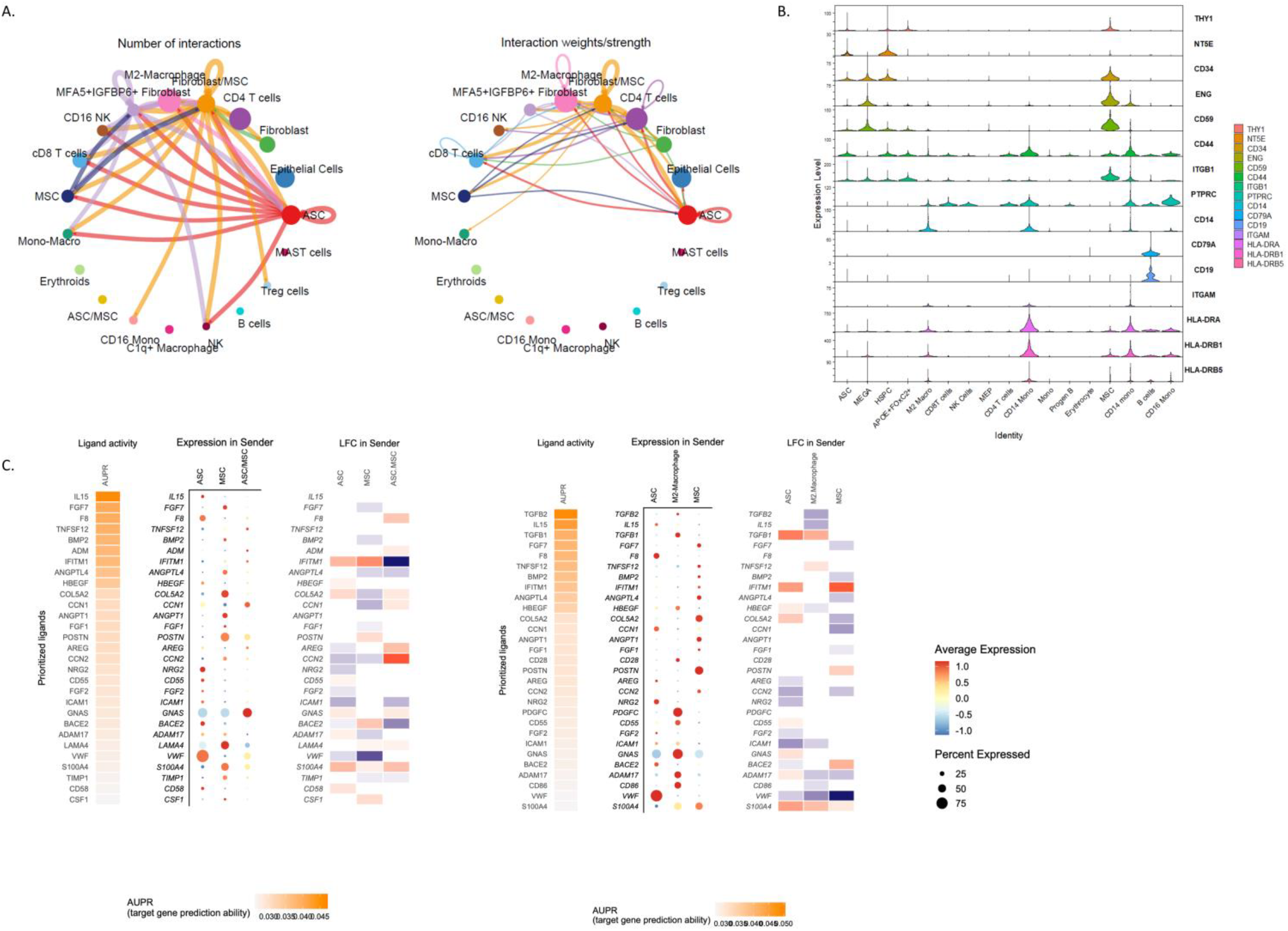
Interaction of Ligand Activity with Gene Expression and Predicted Regulatory Potential in SVF. **A**. Overall Interactions in SVF celltypes, this chord diagram illustrates the comprehensive landscape of cell-to-cell communication within SVF tissue. Each node represents a different cell type, with the connecting edges visualizing the complex web of potential signaling pathways. The diversity of edge thicknesses reflects the varying strengths of these interactions, providing insight into the intricate signaling network that orchestrates cellular functions and responses in SVF. **B**. Violin plot showing the ASC and MSC specific marker genes to help differentiate adipose stromal cells and mesenchymal stromal cells in the SVF tissue niche. **C**. The dot plot represents ligand activity across various prioritized ligands. The size of each dot corresponds to the percentage of cells expressing a given ligand within the sender population, with larger dots indicating a higher percentage. The color intensity reflects the average expression level of each ligand, with warmer colors (red-orange) representing higher expression levels in responders and cooler colors (yellow) representing lower levels in responders.

Expanding on the analysis, a complementary investigation was conducted to examine the role of M2 macrophages as signal senders in SVF, adding another dimension to the predicted cell communication landscape (**Figure 7D**). The dot plot illustrates distinct ligand expression profiles in M2 macrophages compared to ASCs and MSCs, highlighting key differences in signaling activity. Notably, IFITM1 emerges as a significantly upregulated ligand in M2 macrophages, whereas its expression remains relatively lower in the other two cell types. Similarly, HBEGF, TGFB2, and FGF7 demonstrate increased expression in M2 macrophages, reinforcing their potential role in modulating the responder group’s microenvironment. In contrast, ligands such as CD86, BACE2, and VWF are more prominently expressed in ASCs and MSCs, suggesting differential contributions to the signaling network. This extended analysis underscores the nuanced cellular interactions within SVF and refines our understanding of how distinct cell populations contribute to the observed communication dynamics.

## Discussion

Our study provides a comprehensive transcriptomic analysis of Bone Marrow Aspirate Concentrate (BMAC) and Stromal Vascular Fraction (SVF) samples in the context of osteoarthritis (OA) therapy. By analyzing cellular diversity and molecular expression patterns, our purpose was to identify key factors influencing treatment responses. We explored cell composition, gene expression variability, and cell communication networks, building on our previous work with a focus on treatment response variability.

Our findings reveal marked study site-influenced variability in BMAC, with significant differences across five study sites in the United States affecting both cell composition and gene expression. This variability was more pronounced in BMAC than SVF, likely due to the heterogeneity incurred by the technique of needle aspiration from the bone marrow compared with more uniform sampling from subcutaneous fat. Operational variability may be unavoidable in real-world settings, but this finding nevertheless emphasizes the importance of testing and optimizing normalization techniques to enhance the likelihood that observed patterns reflect biological variability rather than procedural artifacts. We applied a range of normalization methods, including Harmony and rPCA, to adjust for site-related effects while preserving biological variability, showing how application of normalization techniques can potentially influence data interpretation.

Overall, there was no significant enrichment of differential gene expression between responders and non-responders after adjusting for the multiple cell types and number of genes considered. Nevertheless, through scITD analysis, we were able to identify shared pathways of co-regulated genes across cell types in both BMAC and SVF. In BMAC, further implementation of a likelihood ratio test provided some evidence for key pathways that larger sample sizes may show are associated with treatment response. These included oxidative phosphorylation^30^, unfolded protein response^31^, and TNFα signaling^32^, suggestive of the initiation of a senescent^33^, pathophysiological environment^34^, related to cellular stress^35^ and immune regulation^36^. Notably, the unfolded protein response, which is linked to endoplasmic reticulum stress, has been implicated in OA^37^, providing a potential mechanistic connection to disease progression. These findings suggest a heightened cellular stress response and metabolic dysregulation in non-responders, possibly contributing to the diminished therapeutic efficacy observed in these patients.

In contrast, SVF samples showed distinct immune regulatory profiles^38,39^, with stromal cells exhibiting the potential to activate pathways related to innate immune responses and T cell activation^38^, while immune cells were enriched in cytokine signaling and antigen presentation pathways. These differences suggest that while BMAC cells may primarily modulate metabolic and inflammatory responses, SVF cells may have more potential to contribute to immune modulation. This dual role of metabolic dysregulation in BMAC and immune regulation^40-42^ in SVF underscores the complexity of OA pathophysiology and highlights the need to tailor cell-based therapies based on these distinct molecular profiles. The pathway distinctions observed between responders and non-responders in BMAC and the immune-modulatory roles of SVF further emphasize the importance of understanding cellular contributions to therapeutic responses in OA^43^.

The reason why the standard differential expression analysis did not reveal any statistically significant genes (after FDR correction) between responders and non-responders in either BMAC or SVF, whereas the LRT showed a strong pattern of enrichment, has to do with the combination of individual variability and study site confounding. The subset of individuals who failed to complete the study included many who dropped out due to non-response, explaining why their baseline profiles appear to be even more deviant from the healthy controls than the included non-responders. We expect that larger sample sizes will confirm the potential markers of non-response, and better address how to avoid study-site differences.

Cell-cell communication analysis further underscored the potential therapeutic roles of MSCs within BMAC and SVF samples. In BMAC, MSCs were identified as major communicators, interacting with various immune cells. Notably, MSCs from responders showed enriched ligand interactions with *THY1*^*44*^, *CD40*, and *IGF1*^*45*^, which are linked to immune modulation and tissue repair, whereas MSCs from non-responders exhibited elevated HLA signaling^46^, indicative of an inflammatory and possibly an autoimmune profile. This suggests that MSC-mediated signaling may be more coordinated in some responders, potentially contributing to treatment efficacy through enhanced immune cell interactions. In SVF, both MSCs and adipose-derived stromal cells (ASCs) demonstrated active communication networks, particularly in responders. Ligands such as *IL15, FGF7*, and *TNFSF12* showed varying expression patterns^47-49^, with *FGF7* upregulated in MSCs and downregulated in ASCs^50^, indicating nuanced differences in cellular interactions that could affect immune responses and tissue repair in OA^51,52^. All of these cell communication inferences are subject to inter-individual variability.

These findings underscore a major limitation of transcriptomic profiling in the context of therapeutic responses, which is that cell therapy outcomes in OA are subject to both patient-specific factors and site-dependent variability. While genomic profiling holds potential for understanding treatment mechanisms, our results indicate that it may be premature to guide OA therapy based solely on molecular profiling, given the high degree of variability. The suggestive pathways identified in this study provide a foundation for future research to clarify these molecular mechanisms and explore their role in patient response, paving the way for more targeted cell-based therapies in OA.

Overall, this study highlights the importance of accounting for site variability in BMAC samples, the distinct roles of MSCs in BMAC and SVF, and the potential of integrating scRNA-seq and cell communication analyses to refine therapeutic strategies. Our findings contribute to a growing understanding of OA’s cellular dynamics and suggest that personalized cell therapy may require a more nuanced approach, considering both molecular signatures and sample-specific factors to improve treatment outcomes.

## Data Availability

All data produced in the present study will be available online once published in a peer-reviewed journal.

## Supplemental Information

Supplemental Information can be found online at [Link will be provided after publishing with peer-reviewed journal]

## Data and Code availability

The datasets used in this manuscript are available with a reviewer link to GSE292453. Will be made public upon publishing this paper.

## Author Contributions

P.C. performed scRNAseq experiments and analyzed and interpreted the data with guidance from G.G. H.S. and L.K. assisted with sample processing and quality control. S.B., K.M. and H.D. managed the MILES trial and conceived the overall experimental design. C.Y., H.D., and K.R. also contributed to data interpretation. All authors reviewed and approved the final manuscript which was initially drafted by P.C. and G.G.

## Acknowledgements

We wish to acknowledge the Cellular Analysis and Cytometry Core and the Molecular Evolution Core facilities at the Parker H. Petit Institute for Bioengineering and Bioscience at the Georgia Institute of Technology for the use of their shared equipment, services, and expertise. Funding for this work was provided by The Billie and Bernie Marcus Foundation to Emory University and the Georgia Institute of Technology, through the Marcus Center for Therapeutic Cell Characterization and Manufacturing (MC3M). K.R. was supported by the Robert A. Milton Endowed Chair. MC3M is also supported by the Georgia Tech Foundation and the Georgia Research Alliance.

