## Supplementary_figures_tables for "Single-Cell Transcriptomics of Multi-Site Cell Therapy in Osteoarthritis: Tissue-Specific Traits and Treatment Correlations"

Supplementary Document- MILES

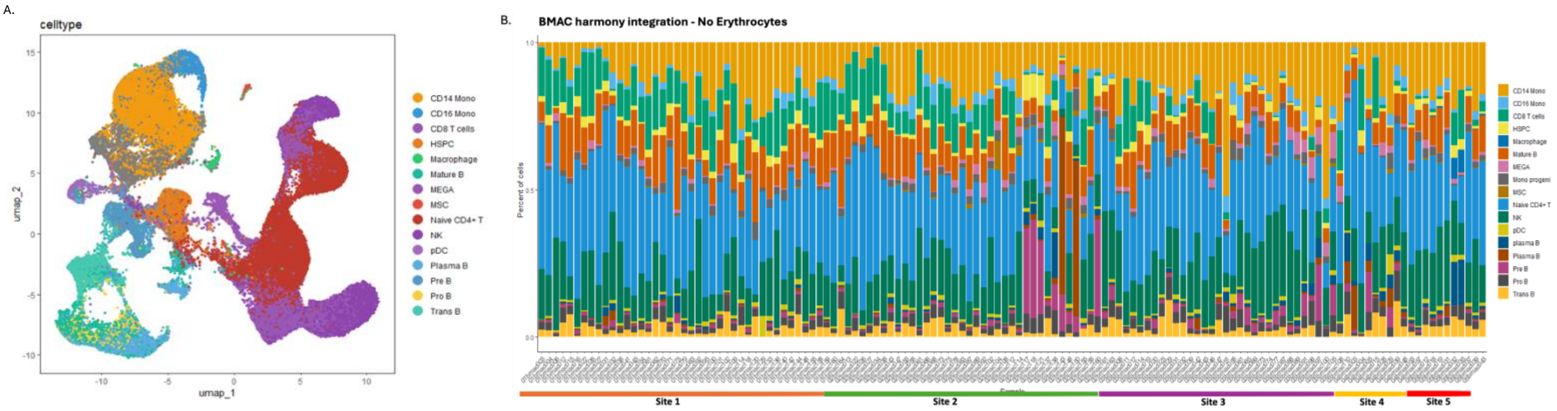


Figure S1: Evaluation of Harmony-Based Integration for Single-Cell Data Normalization
(A) UMAP representation of single-cell transcriptomes from BMAC samples across five clinical sites, integrated using the Harmony algorithm. Cells are colored by annotated cell types based on canonical gene expression markers. Major immune and stromal populations—including CD14+ and CD16+ monocytes, HSPCs, macrophages, various B cell subtypes, MSCs, NK and T cells, and others—were recovered after integration. (B) Stacked bar plot showing the relative proportion of each cell type across all individual samples post-Harmony integration. Samples are grouped by collection site (Sites 1–5), as indicated in the plot.

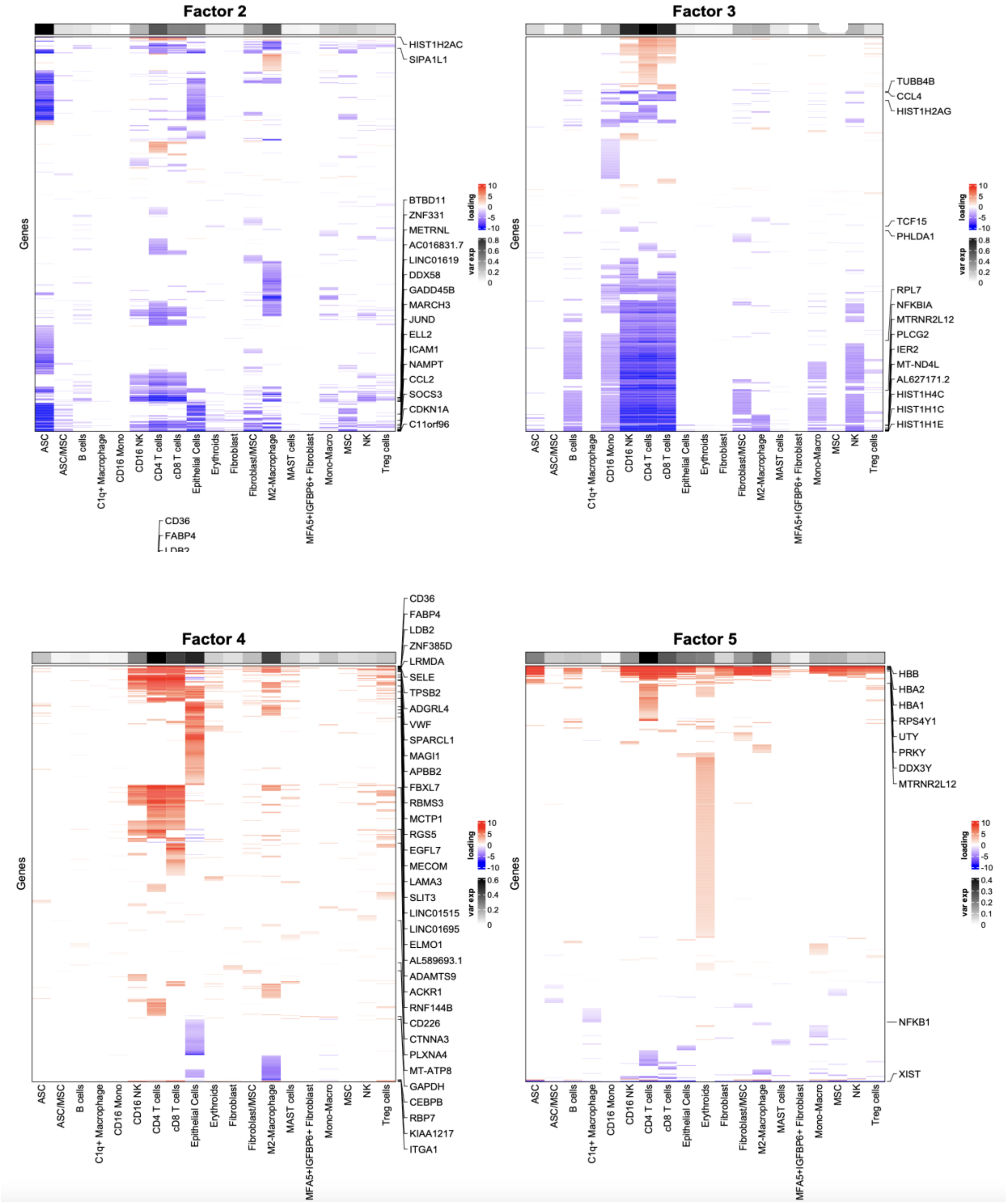


**Figure S2:** Heatmaps illustrating gene expression patterns associated with five distinct factors (Factor 2 to Factor 5) across various cell types. Each heatmap corresponds to a factor derived from a dimensionality reduction analysis, likely non-negative matrix factorization (NMF). The x-axis represents different cell types, while the y-axis lists the genes most strongly associated with each factor. The color scale denotes the relative expression levels, with red indicating higher expression (positive loading values) and blue indicating lower expression (negative loading values). Each factor highlights specific gene expression signatures, suggesting distinct biological processes or cell-type-specific activities. This figure provides insight into the complex regulatory patterns governing gene expression across the examined conditions.

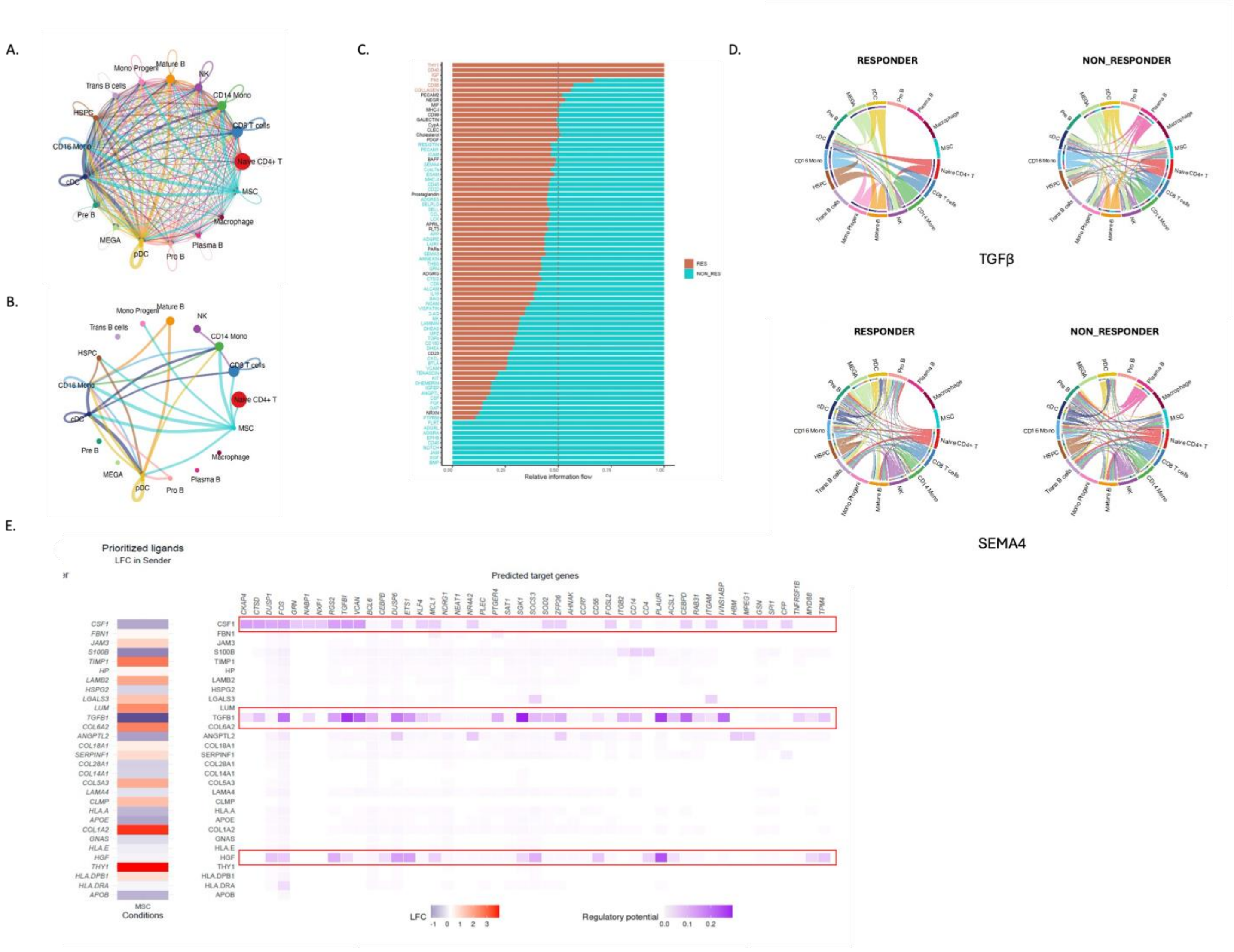


**Figure S3**: **Cell to Cell communications in the tissue BMAC. A.** Overall Interactions in BMAC celltypes, this chord diagram illustrates the comprehensive landscape of cell-to-cell communication within BMAC tissue. Each node represents a different cell type, with the connecting edges visualizing the complex web of potential signaling pathways. The diversity of edge thicknesses reflects the varying strengths of these interactions, providing insight into the intricate signaling network that orchestrates cellular functions and responses in BMAC. **B.** Top 10 communication in BMAC celltypes showing elevated communication from MSCs. This chord plot emphasizes the top 10 communication pathways where mesenchymal stem cells (MSCs) exhibit increased signaling activity. The pronounced edge thickness emanating from MSCs underscores their pivotal role in cell signaling within BMAC, potentially influencing various aspects of tissue behavior and homeostasis. **C.** Relative information flow compared between BMAC responders and non_responders. This comparative plot showing the relative information flow between responder and non-responder cell groups in BMAC. It serves to delineate differences in communication patterns, highlighting how certain cell types may differentially engage in signaling under varying conditions of responsiveness. Such comparative analysis sheds light on the dynamic nature of cellular communication in response to physiological states. **D.** Investigation of the prioritized ligands sending signals from MSCs to other cell types in BMAC and predicted target genes for the selected ligands. The dot plot represents ligand activity across various prioritized ligands. The size of each dot corresponds to the percentage of cells expressing a given ligand within the sender population, with larger dots indicating a higher percentage. The color intensity reflects the average expression level of each ligand, with warmer colors (red-orange) representing higher expression levels and cooler colors (yellow) representing lower levels. Heatmap display of normalized gene expression levels for the same set of ligands in the sender population. Red denotes upregulated genes, and blue denotes downregulated genes, with color intensity reflecting the magnitude of log fold change (LFC). This heatmap provides a focused view of LFC in gene expression for the sender population. Red shades indicate positive LFC, while blue shades indicate negative LFC, again with color intensity corresponding to the degree of fold change. Each column represents a gene predicted to be a target of the corresponding ligand (rows), with the regulatory potential of the interaction depicted by the purple shade's intensity. The darker the shade, the higher the regulatory potential, as measured by the area under the precision-recall curve (AUPR) metric displayed at the bottom, which indicates the ability of ligands to predict their target genes' activity. Color scales for average expression and LFC are included below the heatmaps. Note that specific numeric values and percentages are not indicated on the figure itself and should be referenced in the accompanying text for precise measurements. **E.** Key ligand-receptor pathways like TGFb and SEMA4 show subtle differences in the treatment response groups for Plasma B cells and MSCs.

**Table 1:**

| Celltype | BaselineProp | PropMean.1 | PropMean.2 | PropMean.3 | PropMean.4 | PropMean.5 | Fstatistic | P.Value | FDR |
|---|---|---|---|---|---|---|---|---|---|
| Pre B | 0.03 | 0.023 | 0.068 | 0.05 | 0.055 | 0.03 | 5.4853 | 0.0004 | 0.0068 |
| Plasma B | 0.006 | 0.005 | 0.013 | 0.015 | 0.02 | 0.007 | 2.4526 | 0.0028 | 0.0161 |
| MEGA | 0.01 | 0.006 | 0.016 | 0.022 | 0.008 | 0.011 | 3.3585 | 0.0118 | 0.0401 |
| HSPC | 0.021 | 0.024 | 0.011 | 0.016 | 0.024 | 0.019 | 2.613 | 0.0381 | 0.0926 |
| Mature B | 0.116 | 0.128 | 0.11 | 0.1 | 0.112 | 0.136 | 1.8773 | 0.118 | 0.2408 |
| Trans B cells | 0.032 | 0.038 | 0.033 | 0.034 | 0.04 | 0.056 | 1.8013 | 0.1322 | 0.2048 |
| pDC | 0.008 | 0.009 | 0.008 | 0.008 | 0.008 | 0.013 | 1.261 | 0.2885 | 0.3773 |
| CD16 Mono | 0.015 | 0.015 | 0.015 | 0.019 | 0.017 | 0.013 | 0.9717 | 0.4253 | 0.482 |
| Mono Progeni | 0.036 | 0.034 | 0.04 | 0.039 | 0.037 | 0.037 | 0.4092 | 0.8018 | 0.8018 |

**Table S1:** Cell Proportion analysis Site-based BMAC. This table presents the average proportions of selected cell types across five clinical sites (PropMean.1 to PropMean.5) from single-cell RNA-seq data of bone marrow aspirate concentrate (BMAC) samples. "BaselineProp" indicates the mean proportion of each cell type across all samples. A one-way ANOVA was conducted to assess site-specific variation in cell type proportions. P.Value represents the unadjusted significance level, and FDR is the Benjamini-Hochberg adjusted P-value correcting for multiple comparisons. Cell types with FDR < 0.05 (e.g., Pre B, Plasma B, MEGA) show significant variation across sites. Cell types with FDR ≥ 0.05 are not significantly different across sites.

**Table 2:**

| Celltype | BaselineProp | PropMean.control | PropMean.incmlpt | PropMean.non_responder | PropMean.responder | Fstatistic | P.Value | FDR |
|---|---|---|---|---|---|---|---|---|
| HSPC | 0.021 | 0.016 | 0.027 | 0.016 | 0.023 | 2.9961 | 0.0303 | 0.3819 |
| CD16 Mono | 0.015 | 0.018 | 0.013 | 0.016 | 0.015 | 2.3279 | 0.0774 | 0.3819 |
| Mature B | 0.116 | 0.115 | 0.154 | 0.101 | 0.115 | 2.0322 | 0.1123 | 0.3819 |
| Macrophage | 0.006 | 0.008 | 0.006 | 0.005 | 0.008 | 1.5584 | 0.2025 | 0.4818 |
| CD8 T cells | 0.18 | 0.169 | 0.209 | 0.149 | 0.171 | 1.9994 | 0.3953 | 0.694 |
| Pro B | 0.007 | 0.007 | 0.008 | 0.006 | 0.008 | 0.8823 | 0.449 | 0.694 |
| NK | 0.087 | 0.068 | 0.087 | 0.081 | 0.078 | 0.6691 | 0.5724 | 0.7015 |
| MSC | 0.002 | 0.002 | 0.004 | 0.002 | 0.004 | 0.3392 | 0.8808 | 0.8604 |
| pDC | 0.008 | 0.011 | 0.009 | 0.008 | 0.008 | 0.2176 | 0.8841 | 0.8841 |

**Table S2:** Condition-Based Cell Proportion Analysis Across BMAC Treatment Groups
This table summarizes the average proportions of selected cell types across four treatment conditions: control, incomplete responders, non-responders, and responders. "BaselineProp" represents the mean cell type proportion across all samples. Group-wise means are listed: PropMean.control, PropMean.incmplt (incomplete responders), PropMean.non_responder, PropMean.responder. Statistical differences in cell type proportions across these conditions were assessed using one-way ANOVA. P.Value denotes the unadjusted test significance. FDR is the Benjamini-Hochberg corrected false

discovery rate. No cell types showed statistically significant differences between conditions at FDR < 0.05. However, HSPC displayed nominal significance (P = 0.0303) prior to correction.

**Table 3:**

| Celltype | BaselineProp | PropMean.1 | PropMean.2 | PropMean.3 | PropMean.4 | PropMean.5 | Fstatistic | P.Value | FDR |
|---|---|---|---|---|---|---|---|---|---|
| CD4 T cells | 0.125 | 0.15 | 0.15 | 0.102 | 0.149 | 0.158 | 4.6563 | 0.0015 | 0.2825 |
| Erythroids | 0.029 | 0.026 | 0.036 | 0.024 | 0.011 | 0.011 | 3.4278 | 0.0106 | 0.067 |
| C1q+ Macrophage | 0.019 | 0.022 | 0.018 | 0.017 | 0.012 | 0.012 | 1.9087 | 0.1126 | 0.4279 |
| Treg cells | 0.011 | 0.013 | 0.014 | 0.008 | 0.007 | 0.015 | 1.4063 | 0.2351 | 0.5881 |
| ASC/MSC | 0.019 | 0.018 | 0.014 | 0.005 | 0.022 | 0.009 | 1.1548 | 0.3337 | 0.5881 |
| NK | 0.011 | 0.013 | 0.011 | 0.01 | 0.012 | 0.016 | 1.108 | 0.3555 | 0.5881 |
| MSC | 0.051 | 0.047 | 0.061 | 0.051 | 0.059 | 0.058 | 1.0441 | 0.4024 | 0.5881 |
| B cells | 0.011 | 0.011 | 0.01 | 0.012 | 0.01 | 0.005 | 0.8807 | 0.4774 | 0.6047 |
| CD16 NK | 0.038 | 0.036 | 0.04 | 0.047 | 0.044 | 0.041 | 0.3659 | 0.8326 | 0.9305 |
| cD8 T cells | 0.066 | 0.073 | 0.071 | 0.063 | 0.086 | 0.079 | 1.1209 | 0.9748 | 0.9748 |

**Table S3:** Site-Based Cell Proportion Analysis of SVF Samples. This table presents the average proportions of selected stromal vascular fraction (SVF) cell types across five clinical sites (PropMean.1 to PropMean.5), based on single-cell RNA-seq data. "BaselineProp" indicates the overall mean proportion of each cell type across all samples. A one-way ANOVA was performed to evaluate differences in cell type abundance across sites. P.Value reflects unadjusted significance for site-specific differences. FDR represents the Benjamini-Hochberg adjusted P-value to correct for multiple testing. Although CD4 T cells and Erythroids showed nominal significance (P = 0.0015 and P = 0.0106, respectively), no cell types reached FDR significance (< 0.05), indicating that inter-site differences were not statistically robust after correction.

Table 4:

| Celltype | BaselineProp | PropMean.control | PropMean.incmlpt | PropMean.non_responder | PropMean.responder | Fstatistic | P.Value | FDR |
|---|---|---|---|---|---|---|---|---|
| MAST cells | 0.008 | 0.007 | 0.01 | 0.013 | 0.011 | 2.6205 | 0.0041 | 0.0179 |
| ASC | 0.102 | 0.055 | 0.091 | 0.136 | 0.118 | 2.6904 | 0.0484 | 0.2268 |
| NK | 0.011 | 0.012 | 0.01 | 0.016 | 0.011 | 2.5342 | 0.0595 | 0.2268 |
| Epithelial Cells | 0.107 | 0.12 | 0.154 | 0.111 | 0.127 | 2.0906 | 0.1044 | 0.2558 |
| CD16 Mono | 0.019 | 0.054 | 0.021 | 0.009 | 0.01 | 1.7797 | 0.1505 | 0.3252 |
| Treg cells | 0.011 | 0.012 | 0.01 | 0.015 | 0.012 | 1.608 | 0.1905 | 0.3291 |
| Fibroblast/MSC | 0.019 | 0.1 | 0.061 | 0.094 | 0.089 | 0.9047 | 0.4287 | 0.6231 |
| MSC | 0.051 | 0.054 | 0.053 | 0.006 | 0.052 | 0.4512 | 0.7127 | 0.8489 |
| Mono-Macro | 0.03 | 0.032 | 0.021 | 0.027 | 0.031 | 0.3961 | 0.7594 | 0.8489 |
| Erythroids | 0.029 | 0.025 | 0.029 | 0.026 | 0.023 | 0.0925 | 0.9641 | 0.9641 |

**Table S4:** Condition-Based Cell Proportion Analysis of SVF Samples

This table summarizes the average proportions of selected cell types in SVF (stromal vascular fraction) tissue across four treatment conditions: control, incomplete responders (incmplt), non-responders, and responders. “BaselineProp” indicates the average cell type abundance across all samples. Condition-specific averages are shown in: PropMean.control, PropMean.incmplt, PropMean.non_responder, PropMean.responder. Statistical comparisons were performed using one-way ANOVA. P.Value represents the uncorrected significance for variation across treatment groups. FDR is the Benjamini-Hochberg adjusted false discovery rate to account for multiple testing. MAST cells showed a statistically significant difference across conditions (FDR = 0.0179), suggesting potential condition-specific variation in their abundance. No other cell types reached FDR significance.
